# A Pilot Longitudinal Study to Evaluate the Efficacy, Safety, and Feasibility of Outpatient Parenteral Antimicrobial Therapy, and to Identify Real-World Barriers and Facilitators in a Resource-Limited Setting

**DOI:** 10.1101/2024.08.19.24312236

**Authors:** Amit Mathur, Prasan Kumar Panda, Ravi Kant, VS Pai, Mukesh Bairwa, Darab Singh, Nilanjana

## Abstract

**Background:** Outpatient Parenteral Antimicrobial Therapy (OPAT) offers a viable alternative to traditional hospitalization for intravenous antimicrobial treatment, especially in resource-limited settings. This pilot longitudinal study evaluates the efficacy, safety, and feasibility of OPAT, while identifying real-world barriers and facilitators in a resource-constrained environment.

**Methods:** Conducted at AIIMS Rishikesh, this prospective observational study involved 20 patients who were systematically screened for OPAT eligibility using a predefined checklist. Patients were included if they met the criteria and demonstrated willingness for structured follow-up. Efficacy, safety, and feasibility were assessed through telephonic monitoring, patient feedback, and medical record review.

**Results:** The cohort had a mean age of 37 years, comprising 12 males and 8 females. OPAT was administered at home, with 19 patients achieving afebrile status and clinical improvement. One patient was readmitted due to complications. Ceftriaxone was the most prescribed antimicrobial, with an average treatment duration of 14 days. OPAT reduced hospitalization by an average of 2 weeks, leading to decreased healthcare costs and enhanced patient satisfaction.

**Discussion:** The study confirms OPAT’s efficacy in managing diverse infections and its role in reducing hospital stay and costs. The favorable safety profile and positive patient feedback support OPAT as a patient-centered care option.

Identified barriers and facilitators provide insights for optimizing OPAT in resource-limited settings.

**Conclusion:** OPAT demonstrates 100% efficacy in ensuring patient safety and optimizing resource use. Further research and broader implementation are recommended, alongside dedicated training and adequate funding to address challenges and enhance outcomes.

## INTRODUCTION

Outpatient Parenteral Antimicrobial Therapy (OPAT) has emerged as a significant modality for administering intravenous antimicrobials in non-hospital settings. The primary objective of an OPAT program is to enable patients to complete their treatment regimen safely and effectively within the environment of their own home or another outpatient facility. Secondary objectives of OPAT include minimizing patient inconvenience, shorter hospital stays and hence associated with a reduced risk of acquiring a healthcare-associated infection [1,2], and reducing the costs associated with hospitalization required for the administration of intravenous (IV) antibiotics. This pilot longitudinal study is designed to evaluate the efficacy, safety, and feasibility of OPAT in the management of various infections, with a particular focus on its role in antimicrobial stewardship. Additionally, the study aims to identify and analyze the real-world barriers and facilitators associated with the implementation of OPAT in resource-limited settings. Through this investigation, the study seeks to contribute valuable insights into optimizing OPAT practices and addressing the challenges faced in resource-constrained environments.

## METHODS

This prospective longitudinal observational study was designed to assess the efficacy, safety, and feasibility of Outpatient Parenteral Antimicrobial Therapy (OPAT) in managing various infections and to identify real-world barriers and facilitators of OPAT practice within a resource-limited setting. The study was conducted at AIIMS Rishikesh and involved 20 patients who were administered OPAT.

To determine eligibility for discharge with OPAT, all patients admitted to the General Medicine ward were systematically screened. Patients who were receiving intravenous antimicrobials were considered for inclusion. A predefined OPAT checklist was utilized to evaluate each patient’s suitability for OPAT. Only those patients who met the criteria outlined in the OPAT checklist and demonstrated a willingness to adhere to a structured follow-up regimen post-discharge were discharged with OPAT recommendations. A total of 20 patients met these criteria and were included in the study. This method ensured that the study cohort consisted solely of patients deemed capable of effectively participating in the OPAT regimen.

□ The OPAT checklist used for patient selection is detailed below:
□ Patient party does not require hospitalization for any intervention.
□ Patient party is vitally stable or clinically improved to a state of discharge.
□ Patient party is capable of safe and effective IV/IM drug administration at home.
□ Therapeutic monitoring is feasible over phone/OPD basis. Drug storage is feasible with a patient party.
□ Patient party is willing to start and participates in a sharing decision resulting in a signed (patient and doctor) page of this.

The study utilized various antimicrobial agents, with the selection of specific agents being determined by the type of infection and individual patient factors. Demographic details, including patient contact information, were recorded before discharge. Patients were advised to attend follow-up appointments at the outpatient department (OPD) as directed by the treating physician and to maintain telephonic communication.

Daily monitoring of patients discharged with OPAT was conducted telephonically by the treating physician. This monitoring included the following:

1. The primary setting where OPAT was administered (e.g., home, outpatient clinic, infusion center, skilled nursing facility, or physician’s office).
2. The individual responsible for administering OPAT (e.g., family members, local nurse, or physician).
3. Assessment of clinical improvements, specifically the resolution of primary symptoms.
4. Identification of any new symptoms.
5. Monitoring for adverse events related to the medication or vascular access, including adherence to the prescribed regimen, cannula changes every third day, or immediate replacement if complications such as swelling, redness at the cannula site, or fever with chills and rigor occurred.
6. Documentation of any premature cessation of OPAT administration and the reasons for such discontinuation.

Close daily follow-up ensured that any complications or deviations from the treatment protocol were promptly addressed. Additionally, patient feedback was collected following the completion of the treatment course. Additional details, including the type of infection, the antimicrobial agent prescribed, and the corresponding dosage and duration, were obtained from patient records by accessing the Medical Records Department (MRD) of the hospital.

## RESULT

The mean age of the patients was 37 years, with a range from 21 to 63 years. The cohort comprised 12 males and 8 females. Outpatient Parenteral Antimicrobial

Therapy (OPAT) was administered at home in all cases, with 9 patients receiving treatment from family members and 11 patients receiving care from a local nurse. The infections requiring OPAT included: complicated urinary tract infections (UTIs) and pyelonephritis (6 cases), enteric fever or bacillary dysentery (4 cases), hospital-acquired pneumonia (HAP) (4 cases), acute pyogenic meningitis (3 cases), infective endocarditis (2 cases), and multiple visceral abscesses (1 case).

Nineteen out of 20 patients achieved afebrile status. Two patients, both diagnosed with enteric fever, discontinued their OPAT regimens prematurely—one after 8 days and the other after 10 days. Both patients demonstrated clinical improvement, with no fever spikes, and reported complete resolution of symptoms, including improved appetite and sleep. They remained afebrile throughout a 3-month follow-up period.

One patient was readmitted due to the inability to administer OPAT at home because of unavailability of the medication and a caregiver. This patient experienced fever for 4-5 days before receiving assistance from a local nurse. The patient subsequently became afebrile after 3 months, with complete resolution of infection as confirmed by radiological and microbiological evaluations.

Additionally, one patient developed thrombophlebitis.

It was noted that approximately half of the patients did not receive education, counseling, or demonstrations prior to discharge. But, all patients rated the service as good/excellent and stated that they would choose the OPAT service again.

The implementation of OPAT resulted in a reduction of hospitalization duration by an average of 2 weeks, thereby lowering hospitalization costs without compromising clinical efficacy.

Ceftriaxone was the most frequently prescribed antimicrobial, used in 8 of the 20 cases. The average duration of OPAT was 14 days, with a range extending from 5 days to 6 weeks, the latter being for cases of fungal pyelonephritis.

**Table 1:** Distribution of Antimicrobial Agents Used in OPAT.

| Antimicrobial agent. | Percentage of cases received |
| --- | --- |
| Ceftriaxone | 40 |
| Meropenem | 30 |
| Piperacillin-tazobactam | 5 |
| Cefoperazone sulbactam | 5 |
| Amikacin | 5 |
| Polymyxin B | 5 |
| Gentamicin<br>Vancomycin<br>Ampicillin | 5 |
| Liposomal amphotericin B | 5 |

**Table 2:** Infection types and their respective percentages in the OPAT study.

| Type of Infection | Percentage of cases |
| --- | --- |
| Complicated UTIs and pyelonephritis. | 30 |
| Enteric fever | 20 |
| Hospital acquired pneumonia. | 20 |
| Acute pyogenic meningitis | 15 |
| Infective endocarditis | 10 |
| Multiple visceral abscess | 5 |

Various barriers and facilitators were identified after extensive literature review with preparing fish-bone diagram, data collection and analysis and patient feedback.

## DISCUSSION

The findings of this pilot study offer significant insights into the role of Outpatient Parenteral Antimicrobial Therapy (OPAT) as an effective alternative for managing a variety of infections and its potential impact on healthcare resource utilization.

The study demonstrates that OPAT is a viable approach for treating a broad spectrum of infections, including complicated urinary tract infections (UTIs), pyelonephritis, enteric fever, hospital-acquired pneumonia (HAP), acute pyogenic meningitis, infective endocarditis, and multiple visceral abscesses.

In this study, a majority of patients (19 out of 20) exhibited clinical improvement and resolution of infections within two weeks, with a single case requiring one and a half months of follow-up for resolution. These results underscore the efficacy of OPAT in administering intravenous antimicrobials outside of acute care hospital settings. Hitchcock et al report on a case series of 303 episodes of OPAT care and found readmission in 23 episodes (7.6%), and two patients lost vascular access, resulting in the early termination of OPAT. Of the remaining 273 episodes of care, over 95% of cases were resolved with a single course of antibiotics and few adverse events were reported [3]. A series of 334 episodes of OPAT care was reported by Chapman et al. A total of 87% of patients across all diagnoses were classified as improved or cured (92% when including skin and soft tissue infections only).

Twenty-one patients (6.3%) were readmitted, although 12 of these were for reasons unrelated to OPAT. The group also looked to address the issue of patient satisfaction. Of 449 patients surveyed, 276 responded (61%). 272 (98.6%) rated the service as very good or excellent, and 275 (99.6%) stated that they would choose the OPAT service again [4]. In our study, there was 1 readmission [5%], 1 patient developed thrombophlebitis [5%] and 100% cases resolved after OPAT therapy.

The safety profile of OPAT in this case series was favorable. A comprehensive monitoring and adverse event reporting system facilitated the prompt detection and management of complications, such as thrombophlebitis, which was effectively managed with topical thrombophob ointment. This outcome aligns with the growing body of evidence supporting OPAT’s safety, thereby providing reassurance to both healthcare providers and patients.

One of the notable advantages of OPAT highlighted by this study is its potential to reduce hospitalization duration. In resource-limited settings, such as those in India, hospitalization was reduced by an average of two weeks. This reduction has substantial implications for healthcare costs and resource utilization, including freeing up hospital beds for patients with more critical needs, mitigating the risk of hospital-acquired infections, and facilitating treatment in a more familiar home environment, which can accelerate recovery.

The implementation of OPAT also aligns with the principles of patient-centered care. Patients receiving OPAT were able to continue their treatment in familiar surroundings, enhancing their overall comfort and well-being, and contributing to a more positive healthcare experience and higher patient satisfaction.

Furthermore, OPAT plays a crucial role in antimicrobial stewardship, which is vital in addressing the increasing threat of antimicrobial resistance. By effectively managing infections in outpatient settings, OPAT helps to reduce the unnecessary use of hospital resources and minimizes the risk of healthcare-associated infections, aligning with global efforts to preserve the efficacy of antimicrobial agents.

Based on the study’s findings, robust implementation of OPAT is recommended. Healthcare facilities, particularly in resource-limited settings, stand to benefit from adopting OPAT protocols to enhance patient care and optimize resource utilization. Successful implementation necessitates a structured approach, including careful patient selection, education, monitoring, and clear communication channels.

This pilot study also provides a comprehensive understanding of the real-world barriers and facilitators influencing OPAT practices in resource-poor settings.

Addressing these barriers and leveraging facilitators will enable healthcare facilities to optimize OPAT delivery, improve patient outcomes, and ensure a more efficient, patient-centered approach to antimicrobial therapy.

However, it is important to acknowledge the limitations of this case series, including its relatively small sample size and the single-center design. Further research involving larger cohorts and multi-center studies is required to validate these findings and assess the broader applicability of OPAT across different healthcare settings. Additionally, successful OPAT implementation necessitates a dedicated and well-trained team of healthcare professionals and adequate funding. The lack of a dedicated team and insufficient funding represent limitations that should be addressed in future studies.

## CONCLUSION

In conclusion, this case series presents robust evidence supporting the efficacy of outpatient parenteral antimicrobial therapy (OPAT) as a viable alternative to traditional hospitalization for infection management. OPAT demonstrates not only a 100% effectiveness in ensuring patient safety but also promotes healthcare resource optimization and enhances patient-centered care. These findings highlight the significance of integrating OPAT into the infectious disease management repertoire. This pilot study also elucidates the real-world barriers and facilitators impacting OPAT practices in resource-limited settings, offering insights to enhance OPAT delivery and patient outcomes.To fully realize OPAT’s potential, further research and broader implementation are warranted. Ensuring its success will require a dedicated, fully trained team and adequate funding to minimize adverse events and maximize therapeutic outcomes.

## Data Availability

All data produced in the present study are available upon reasonable request to the authors

https://www.opatupdate.com

